# The Limits of Prompt Engineering in Medical Problem-Solving: A Comparative Analysis with ChatGPT on calculation based USMLE Medical Questions

**DOI:** 10.1101/2023.08.06.23293710

**Authors:** Dhavalkumar Patel, Ganesh Raut, Eyal Zimlichman, Satya Narayan Cheetirala, Girish Nadkarni, Benjamin S Glicksberg, Robert Freeman, Prem Timsina, Eyal Klang

**Affiliations:** Mount Sinai Health System; Hospital Management, Sheba Medical Center, Affiliated to Tel-Aviv University; ARC Innovation Center, Sheba Medical Center, Affiliated to Tel-Aviv University; The Charles Bronfman Institute of Personalized Medicine, Icahn School of Medicine at Mount Sinai, New York, New York, USA

**Keywords:** Large Language Models, Prompt Engineering, GPT-3.5, GPT-4, Medical Problem-Solving, USMLE, Chain of Thoughts, Modified Chain of Thoughts

## Abstract

**Background:** Prompt engineering significantly improves the performance of Large Language Models (LLMs), including GPT-3.5 and GPT-4. However, its utilization remains largely uncharted in the medical field.

**Objective:** This research aimed to assess the influence of different prompt engineering strategies on ChatGPT (GPT-3.5) in solving medical problems, specifically focusing on medical calculations and clinical scenarios.

**Design:** We utilized three different prompting strategies—direct prompting, the chain of thoughts (CoT), and a modified CoT method—across two sets of USMLE-style questions.

**Setting:** The experiment was conducted using a 1000-question dataset, generated by GPT-4 with a specialized prompt, and a secondary analysis with 95 actual USMLE Step 1 questions.

**Measurements:** Model performance was assessed based on accuracy in answering medical calculation and clinical scenario questions across varying difficulty levels and medical subjects.

**Results:** Direct prompting demonstrated non-inferior accuracy compared to the CoT and modified CoT methods in both question categories. This trend remained consistent regardless of difficulty level or subject matter in the GPT-4-generated dataset and USMLE Step 1 sample questions.

**Limitations:** The study evaluated GPT-3.5 for answering and GPT 4 for question generation, limiting generalizability.

**Conclusion:** Our findings indicate that while prompt engineering can facilitate question generation, as exemplified by GPT-4, it does not necessarily improve model performance in answering medical calculation or clinical scenario questions. This suggests that the ChatGPT model is already effectively optimized for such tasks. Additionally, this finding simplifies the use of such models in healthcare settings, allowing practitioners to interact effectively with tools like ChatGPT without the need for complex prompt engineering, potentially encouraging wider adoption in clinical practice for problem-solving, patient care, and continuous learning.

## Introduction

Prompt engineering is fast becoming a crucial tool in maximizing the performance of large language models (LLMs) like ChatGPT [1]. As a rapidly growing field within natural language processing (NLP), it shapes the way we interact with LLMs, including GPT-3.5 and GPT-4 [1-2]. Yet, when it comes to medical problem-solving tasks, the full capacity of prompt engineering remains largely uncharted territory [3,4].

Past studies have highlighted the potential of ChatGPT (GPT-3.5), which managed to score approximately 60% on the USMLE [3]. Surprisingly, the more advanced GPT-4 achieved an impressive 87% [4]. However, it was found that the application of the “chain of thoughts” (CoT) prompt engineering technique, which essentially instructs the model to “reason step by step,” did not lead to any significant improvement in GPT-4’s performance [5-6]. This could be due to the nature of the USMLE exam, which does not heavily rely on mathematical reasoning.

Outside of medicine, the effectiveness of prompt engineering has been demonstrated across a range of tasks [1-2], suggesting its potential for specific applications within the medical field. In particular, tasks that involve calculations could significantly benefit from specialized prompting strategies.

Despite its promise, there are few studies focusing on prompt engineering within the medical field listed on PubMed. This leaves many questions about the efficacy of different prompting strategies for medical problem-solving tasks unanswered.

We set out to address this knowledge gap in this study. We evaluated the performance of GPT-3.5 on calculation and non-calculation type questions in the USMLE Step 1 format. We compared the efficacy of three different prompting strategies: direct prompt, CoT, and a modified CoT. To ensure a fair comparison, the questions were generated by GPT-4, and all three methods were tested on a sample set from the actual USMLE Step 1 exam.

Our objective was to assess whether prompt engineering could enhance the performance of ChatGPT in calculation and clinical medical problem-solving tasks.

## Methods

### Study Design

We examined the performance of the OpenAI language model, ChatGPT (GPT-3.5-turbo) [7], across multiple choice question (MCQ) datasets using three types of prompts: direct prompt, chain of thoughts (CoT), and a modified CoT. We included a sample USMLE Step 1 questionnaire [8], excluding image-based questions, which resulted in 95 remaining questions. We further supplemented this with two additional MCQ datasets generated by the GPT-4 model, one focused on calculations and the other on clinical vignettes.

### Question Generation

With GPT-4, we created 1000 USMLE-style questions [9], dividing them equally into two categories: calculation (n=500) and non-calculation (n=500). Non-calculation questions encompassed diagnosis based on symptoms, treatment choices, lab results interpretation, disease progression, pathophysiology, public health, and preventive medicine questions. Calculation questions involved more quantitative tasks like drug dosage calculations, clinical score computations, diagnostic test calculations, and statistical data interpretation.

Each question was assigned a difficulty level of easy, medium, or hard by GPT-4. The questions spanned 19 different clinical fields, including Internal Medicine, Pediatrics, Psychiatry, Surgery, among others.

For generating questions, the prompt was:

*Dear GPT-4, we are conducting research on prompt engineering and your help is needed to generate a high-quality medical question. This question should meet the following criteria:*

- *It should be similar to those found in the USMLE Step 1 examination*.
- *It should be in the field of {clinical_field}*.
- *It should be a {broad_type} type question, specifically focusing on {subtype}*.
- *Its difficulty level should be {difficulty}*.
- *It should be clear, concise, and directly relevant to the specified medical field*.
- *The correct answer should be unambiguous*.

~~~
*{
      “Question (+ choices)”:* …,
      *“Answer (must be only the single letter of the choice answer)”:*
…
*}*
~~~

Where *clinical_field* was one of 19 fields (internal medicine, surgery, etc.), *broad_type* was either calculation or non-calculation. *Subtypes* were Drug dosage calculations, Clinical score calculations, Diagnostic test calculations, Statistical data interpretation for calculation questions, and Diagnosis based on symptoms, Treatment selection, Interpretation of lab results, Disease progression and prognosis, Pathophysiology questions, and Public health and preventive medicine questions for non-calculation type questions. *Difficulty level* was easy, medium, or hard.

#### Question answering - prompt Engineering

To query GPT-3.5, we used three prompting strategies. The “direct prompt” strategy simply instructed the model to “answer the question.” The “CoT” strategy guided the model to “reason step by step and answer the question.” The “modified CoT” strategy directed the model to “read the problem carefully, break it down, devise a strategy for solving it, check each step for accuracy, and clearly and concisely convey your reasoning leading to your final answer.” This approach sought to mimic human problem-solving behavior, with the aim to enhance the model’s reasoning ability while promoting clarity and precision in its responses.

All prompts were submitted using openAI API with the following format, using default temperature (0.5) and max token length of 700:

~~~
      *{
      “role”: “system”*,
      *“content”: “You are a knowledgeable medical assistant*.*”
      }*,
      *{
      “role”: “user”*,
      *“content”: “prompt”*
~~~

where *‘prompt’* corresponded to:

*question + direct / CoT / modified CoT prompt*.

Where the direct prompt was:

*“Please answer the question”*

The CoT prompt was:

*“Please reason step by step and answer the question”*

The modified CoT prompt was:

*“Please read the problem carefully, break it down into manageable parts. Devise a strategy for solving it. Double-check each step for accuracy. Convey your reasoning clearly and concisely, leading to your final answer*.*”*

### Evaluation

The primary measure of the evaluation was the accuracy of GPT-3.5’s responses as determined via the OpenAI API. This evaluation was also performed on the USMLE Step 1 question sample for comparison.

Additional sub-Analyses were conducted based on question difficulty, type, and medical field to gain a comprehensive understanding of chatGPT performance.

### Statistical Analysis

Statistical analyses were executed using Python version 3.9.16. We used the Chi-square test to examine the correlation between prompt types and response accuracy. A p-value of less than 0.05 was considered statistically significant.

## Results

The results of our study are outlined in **Table 1**. We assessed the performance of GPT-3.5 using three prompting strategies: direct prompt, chain of thoughts (CoT), and a new modified CoT. The performance was evaluated using three different sets of questions: a sample of USMLE Step 1, a set of clinical questions generated by GPT-4, and a set of calculation-oriented questions generated by GPT-4.

**Table 1:** Comparison of using three prompting strategies in chatGPT for USMLE step 1 sample, GPT-4 generated clinical non-calculation questions, and GPT-4 generated calculation medical questions.

|  | ChatGPT<br>direct prompt | ChatGPT<br>CoT | ChatGPT<br>Modified CoT | P value |
| --- | --- | --- | --- | --- |
| **USMLE step 1**<br>**sample** | 54 / 95<br>(61.7%) | 59 / 95<br>(62.8%) | 58 / 95<br>(57.4%) | 0.734 |
| **GPT-4**<br>**Generated clinical** | 270 / 500<br>(54.0%) | 274 / 500<br>(54.8%) | 257 / 500<br>(51.4%) | 0.530 |
| **GPT-4 Generated**<br>**calculation** | 397 / 500<br>(79.4%) | 398 / 500<br>(79.6%) | 386 / 500<br>(77.2%) | 0.589 |

For calculation questions, non-calculation clinical questions, and the USMLE Step 1 sample questions, there was no significant difference in performance between the direct prompt, CoT, and Modified CoT methods (**Table 1**).

We performed sub-analyses on GPT-3.5’s performance, dividing the tasks into calculation and clinical questions, further dissecting these based on difficulty and subtype.

For calculation questions, no specific prompting strategy demonstrated significant superiority over the others, irrespective of task complexity. For example, at the easy difficulty level, the accuracy rates of the three prompt strategies were quite comparable: Direct Prompt (64.2%), CoT (63.1%), and Modified CoT (61.9%) (**Figure 2**). By question subtype, no specific strategy showed a clear advantage (**Figure 3**). p-values were consistently greater than 0.05 for all sub-analyses, suggesting no statistically significant differences.

**Figure 1:**
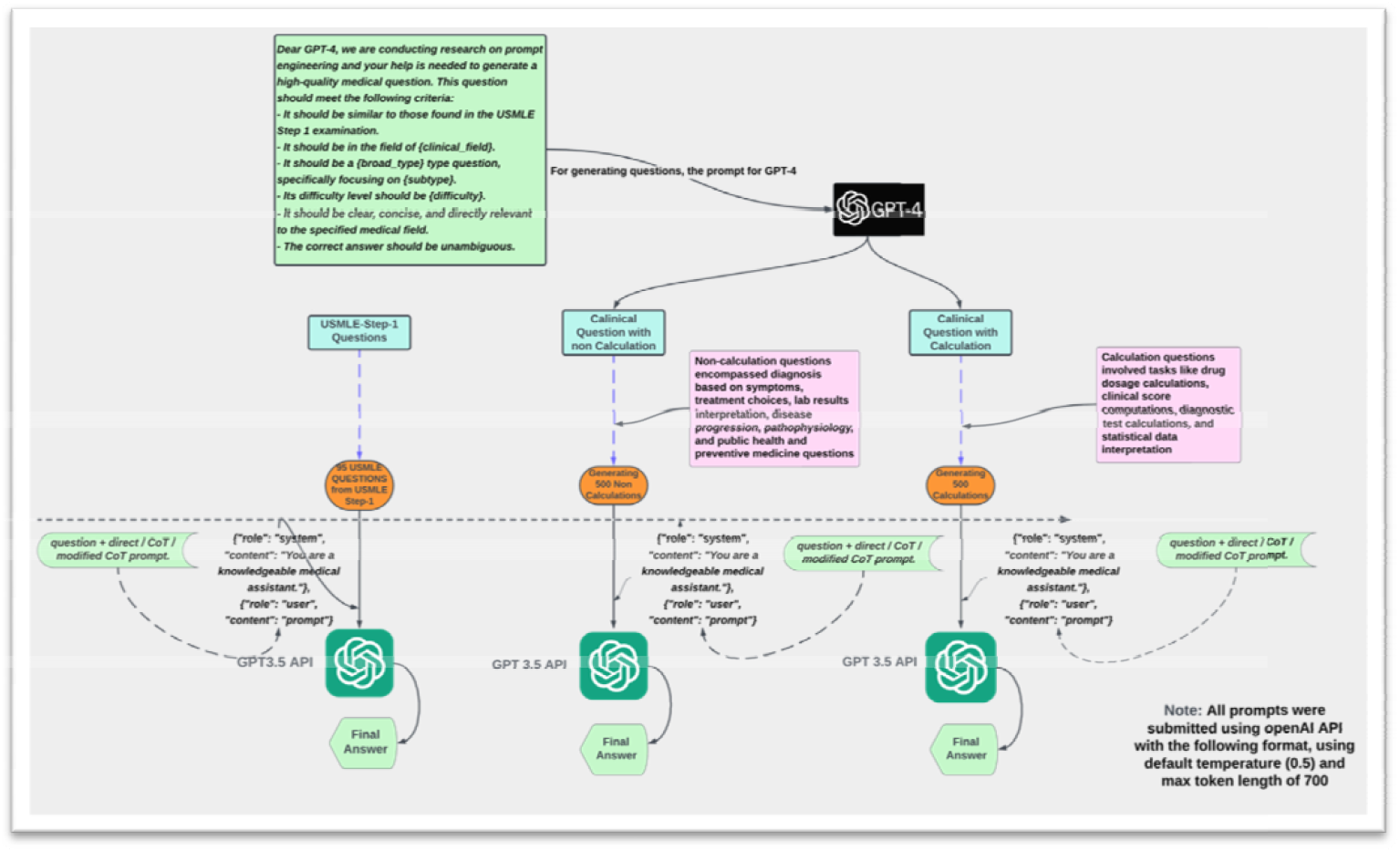
The figure illustrates a multi-step process in which GPT-4 generates USMLE-style medical 100 questions with calculation and non-calculation, and GPT-3.5-turbo answers them using three prompting strategies direct, COT, and Modified COT. The generated questions span 19 clinical fields and various medical topics, and the model’s answers aim to mimic human problem-solving behavior, enhancing reasoning ability and clarity in its responses.

**Figure 2:**
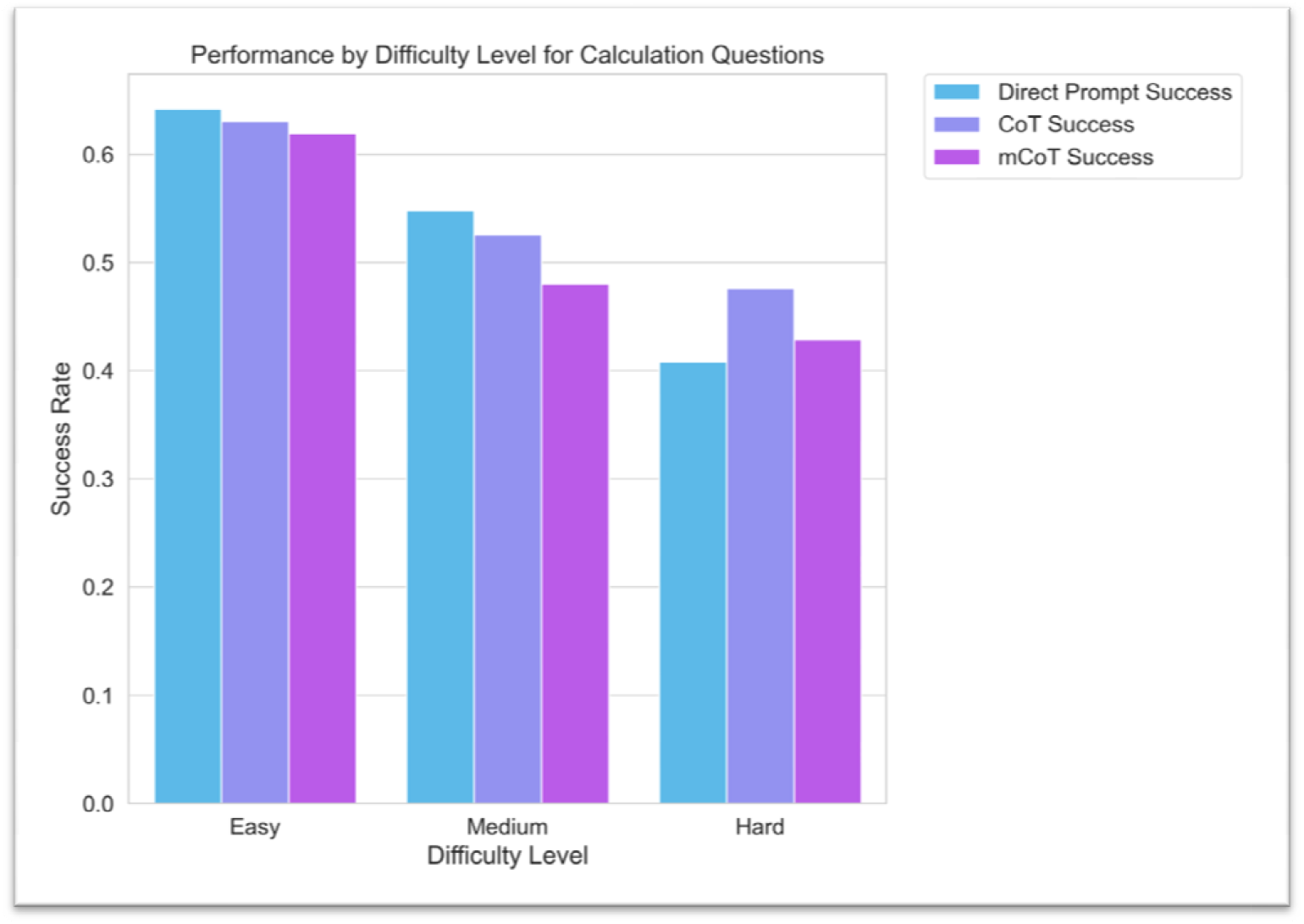
Bar graph representing the success rates of different prompting strategies (‘Direct Prompt’, ‘CoT’, and ‘Modified CoT’) for ‘calculation’ type questions across different difficulty levels (‘Easy’, ‘Medium’, ‘Hard’). Each bar corresponds to the average success rate for the respective prompting strategy and difficulty level.

**Figure 3:**
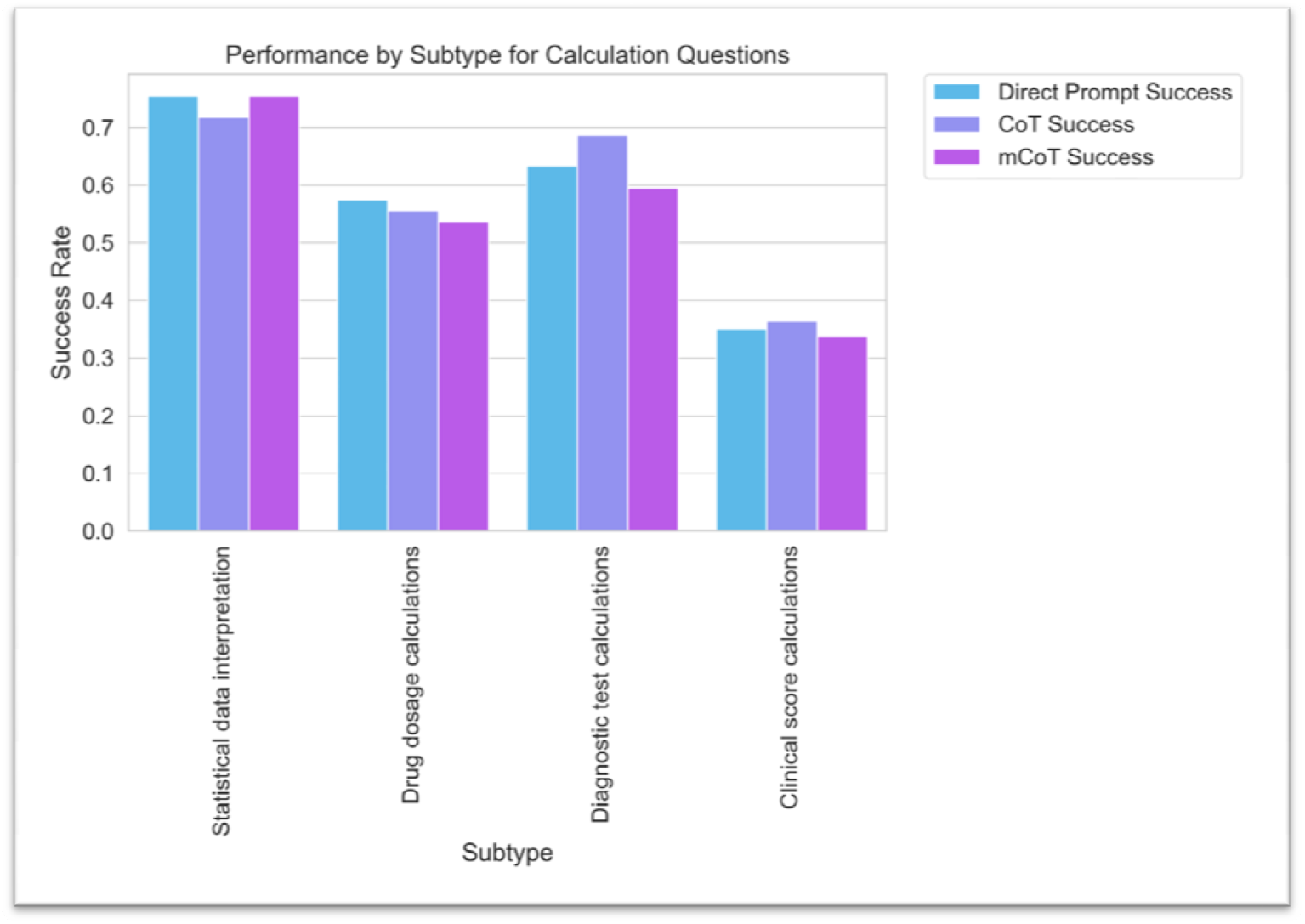
Bar graph showing the success rates of various prompting strategies (‘Direct Prompt’, ‘CoT’, and ‘Modified CoT’) for different ‘calculation’ question subtypes. Each bar represents the average success rate of a particular prompting strategy for a specific subtype.

Similarly, in clinical questions, no single strategy demonstrated clear superiority across differe difficulty levels or question subtypes. For instance, in the diagnosis based on symptoms subtype, the success rates were very similar for all strategies, with Direct Prompt (88.9%), CoT (90.1%), and Modified CoT (90.1%) (**Figures 4 and 5**). These trends remained consistent across other subtypes, such as disease progression and prognosis, interpretation of lab results, and treatment selection, with p-values suggesting no significant difference between the strategies.

**Figure 4:**
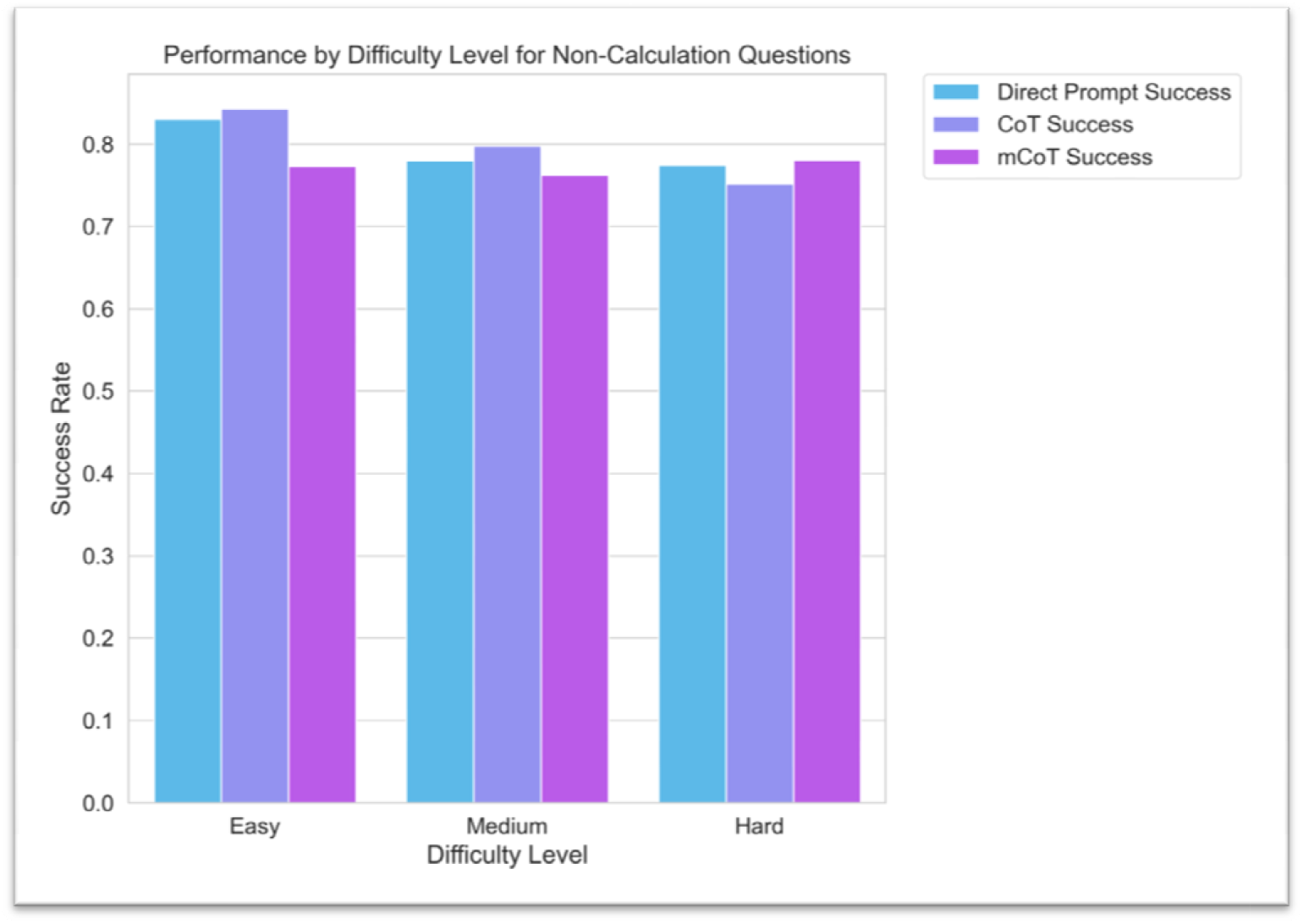
Bar graph depicting the success rates of the three prompting strategies (‘Direct Prompt’, ‘CoT’, an ‘Modified CoT’) for ‘non-calculation’ type questions across different difficulty levels (‘Easy’, ‘Medium’, ‘Hard’). Eac bar corresponds to the average success rate for a specific prompting strategy and difficulty level.

**Figure 5:**
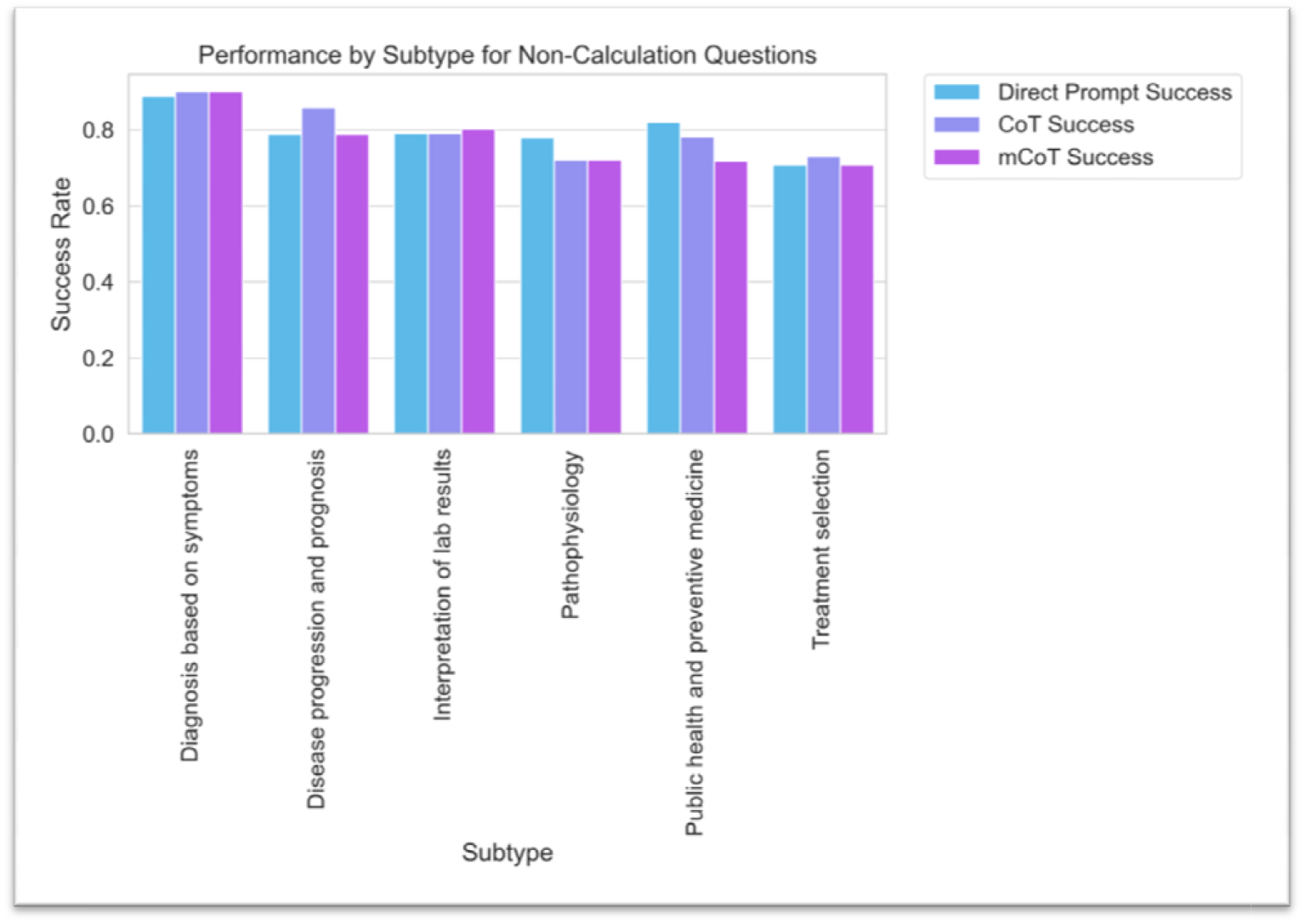
Bar graph illustrating the success rates of different prompting strategies (‘Direct Prompt’, ‘CoT’, and ‘Modified CoT’) for various ‘non-calculation’ question subtypes. Each bar represents the average success rate for a certain prompting strategy for a given subtype.

Overall, these analyses suggest that while individual prompt strategies may have slight variances in success rates for specific tasks, the differences are not statistically significant, indicating that all prompt strategies perform comparably in both calculation and clinical questions, regardless of the difficulty level or subtype.

Upon examining the AI model’s performance across various medical specialties (**Figure 6**), we observed no specific pattern tied to the type of prompting strategy employed, with all p-values much higher than 0.05. However, a distinct trend emerged when looking at the overall performance. Dermatology displayed the highest success rates across all strategies, with an average rate around 79.5%. Conversely, Anesthesiology recorded the lowest success rates, averaging around 49.4%.

**Figure 6:**
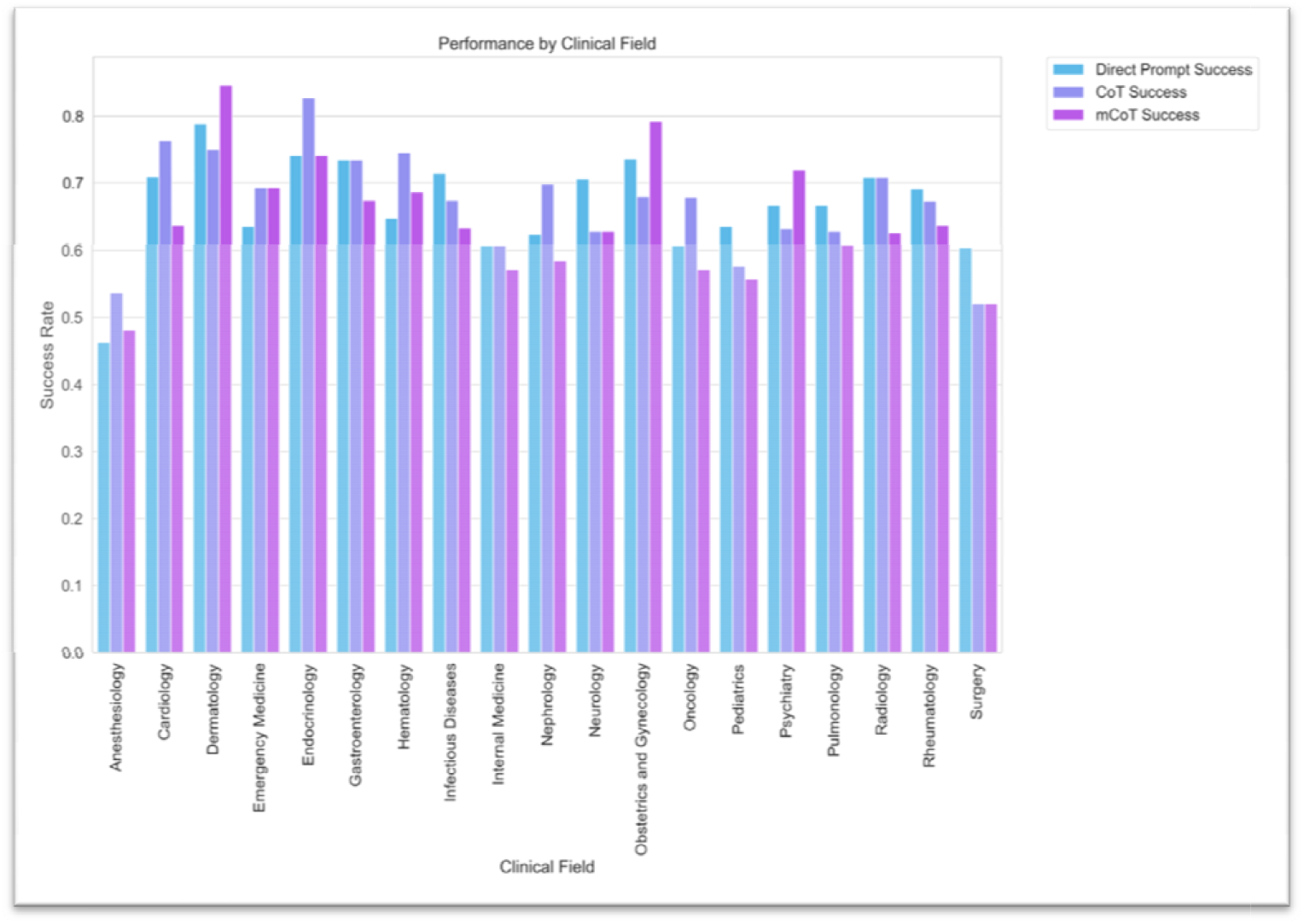
Bar graph detailing the success rates of the three prompting strategies (‘Direct Prompt’, ‘CoT’, and ‘Modified CoT’) across diverse medical fields. Each bar denotes the average success rate for a specific promptin strategy within a particular field.

## Discussion

Our study reveals some surprising and intriguing findings regarding the role of prompt engineering in harnessing the performance of ChatGPT for medical problem-solving tasks. We evaluated three different prompt methods: direct prompt, chain of thoughts (CoT), and a modified version of CoT. Contrary to our initial expectations, we found no significant differences in the performance of these strategies, neither in medical calculation nor non-calculation questions.

In terms of performance, all prompt methods achieved a high percentage of correct answers on medical multiple-choice questions. We chose to test ChatGPT on problem solving and not GPT-4. While the raw computational power of GPT-4 is indisputable, its high cost makes it less feasible for widespread use. In contrast, ChatGPT is not only freely available on the web, but it is also about 30 times more cost-effective when utilized via the OpenAI API [10], making it an accessible and economical option for many real-world applications.

Our results underline the complexity of prompt engineering in relation to medical problem-solving [9]. Despite the uniform effectiveness of the strategies we employed, it is crucial to remember that these are just a few among many potential approaches. We did not, for instance, investigate other techniques like the self-consistency CoT [11], which uses majority voting from multiple CoT attempts, or the “few-shot learning” method [12]. Each of these strategies carries its own strengths and limitations, and further research is needed to understand their implications fully.

The training process of ChatGPT, executed by OpenAI, largely remains a black box. The intricate details about the strategies employed during the training phase or at inference time are not entirely disclosed. However, it is reasonable to assume that OpenAI, well-versed in the landscape of AI and m achine l earning, would incorporate established prompt engineering strategies. In fact, the non-differential performance of our distinct prompt techniques might be a testament to this. The core principles of these strategies may already be integrated into ChatGPT’s functioning, either during training or inference, rendering explicit implementations of prompt engineering techniques, such as CoT, redundant.

The growing sophistication of Large Language Models like ChatGPT signals a future where these AI tools become an integral part of a physician’s professional life. Therefore, it is vital to not only study these models but also incorporate their understanding in medical education. In our study, it was encouraging to find that a simple direct approach yielded comparable results to more elaborate prompting strategies. This finding simplifies the use of these models, as healthcare professionals can effectively interact with ChatGPT and similar tools without needing to master complex prompt engineering techniques. Consequently, this promotes their widespread adoption in clinical practice, where they can assist in medical problem-solving, patient care, and even continuous medical learning.

Nonetheless, our study is not without limitations. Firstly, our findings are drawn from USMLE-style questions, potentially limiting their applicability to other question types or domains. Secondly, our research focused exclusively on ChatGPT. Different LLMs might react differently to these prompt engineering strategies. Thirdly, we did not account for possible variations in the implementation of prompt strategies, which could potentially influence the outcomes. Lastly, by categorizing questions into ‘calculation’ and ‘non-calculation’ types, we may have overlooked the intricate mix of elements often present in medical problem-solving.

In conclusion, the study results suggest that while prompt engineering can be useful for generating datasets, like the questions generated by GPT-4, it does not necessarily enhance model performance when answering calculation or clinical medical type questions. This suggests that the ChatGPT model is already well-optimized for such tasks.

## Supporting information

Prompt Eng Question and Answer -Supplementari Data

## Data Availability

All data produced in the present study are available upon reasonable request to the authors

